# Cervical Stiffness in Mid-Pregnancy: A Predictive Tool Using a Novel Aspiration Device for Spontaneous Preterm Birth Risk Assessment

**DOI:** 10.64898/2025.12.01.25341218

**Authors:** Irene Hösli, Gundula Hebisch, Michael Bajka, Katharina Quack Lötscher, Alice Winkler, Leonhard Schäffer, Begoña Martinez de Tejada, Monya Todesco Bernasconi, Tina Fischer, Vincent Uerlings, Jute Richter, Stephanie Von Orelli, Markus Kuther, Elke Prentl, Ursula Fellmann, Alexander Krafft, Lea Köchli, Sabrina Badir, David Scheiner

## Abstract

**Background:** Preterm birth is a major prenatal care problem. Predicting who will deliver prematurely is particularly challenging in women who do not present known risk factors. Abnormal cervical softening can contribute to a higher risk for spontaneous preterm delivery; a clinically deployable technique to accurately measure cervical stiffness could improve identification of women at risk.

**Objective:** This study aimed at evaluating differences in cervical stiffness measured mid-pregnancy with the aspiration-based Pregnolia System between largely unselected pregnant women who ultimately delivered at term and the ones who had a spontaneous preterm birth; and to evaluate the performance of stiffness in predicting spontaneous preterm birth.

**Study Design:** This was a cross-sectional, prospective, cohort study of pregnant women presenting at their mid pregnancy consultation (gestational age 18^0/7^ – 22^0/7^ weeks) conducted at 13 centers. Cervical stiffness, assessed as Cervical Stiffness Index (CSI), was measured with an aspiration-based device placed on the anterior lip of the cervix during a speculum examination. Stiffness was also determined through the maximum cervical compressibility ratio (Cervical Consistency Index, CCI) and cervical length was measured transvaginally. Pain or discomfort scoring (from 0 – no pain to 10 – maximum possible pain), and delivery information were also recorded.

**Results:** Data from 1002 women were analyzed, of which 990 were singleton pregnancies. Preterm birth rate (5.2%) in the whole dataset was considerably lower than expected, with 25 spontaneous among a total of 52 preterm births. A significant difference in mean CSI between singleton women delivering at term (median=71 mbar, IQR=(51,97), n=948) and women delivering spontaneously prematurely before 37 weeks (65 mbar (42,70), n=19) was found (p=0.039). Instead, the difference in cervical length and CCI was not significant. Women who had no prior vaginal delivery had a cervix on average 15% stiffer (75 mbar (55,103), n=590) than those with a previous vaginal delivery (64 mbar (45, 85), n=358) (p<0.001). In the sub-population without history of vaginal delivery, the difference in CSI between women with a term birth (75 mbar (55,103)) and women with a spontaneous preterm birth (59 mbar (42,70), n=15) was more significant (p=0.007). Mean discomfort score for CSI assessment was 0.51±1.32.

**Conclusion:** Women with singleton pregnancies delivering spontaneously preterm had a significantly lower cervical stiffness than those delivering at term. This difference is even more significant in a sub-population without history of vaginal delivery. Women with a term birth and no history of vaginal delivery had a significantly stiffer cervix than women with a history of vaginal delivery, suggesting that mechanical properties of the cervix are softened by prior vaginal birth. Quantitative measurement of cervical stiffness with the aspiration-based Pregnolia System proved robust for objective measurement of cervical softness during pregnancy, with a very low discomfort score.

## Introduction

Premature birth (PTB) defined as delivery before 37 weeks of gestation is a global, yet unsolved, health issue. With more than 13 million PTBs every year, it is the leading cause of neonatal and infant mortality globally. In the last decade, despite all the efforts in the field, PTB rates have remained stable (9.8% in 2010 and 9.9% in 2020^1^). Prevention of spontaneous PTB (sPTB) remains challenging as both prediction methods and therapies are currently limited^2,3^.

The current standard for PTB risk assessment is to measure cervical length (CL) using transvaginal ultrasound. A CL <25 mm at mid-gestation is considered to increase the risk of PTB in otherwise asymptomatic women. However, this method has only limited predictive capability, as most of the women delivering preterm do not present a short cervix^4–6^.

Cervical softening has long been considered an important descriptive parameter of cervical status throughout pregnancy. Recently, due to technological advances^7,8^, it has regained consideration as having potential to improve sPTB risk assessment^9^. Ultrasound- and aspiration-based tools are being developed to accurately measure the properties of the cervix and objectively quantify its stiffness. Despite the still low number of studies, early results indicate that it performs significantly better than CL in predicting PTB^10–20^.

The Pregnolia System is an aspiration-based CE-certified medical device quantifying cervical stiffness by measuring the amount of vacuum required to gently displace cervical tissue into a probe tip by a fixed distance. This value, called Cervical Stiffness Index (CSI), is a proxy parameter for stiffness: the stiffer the tissue, the higher the pressure. The device has been used in several studies in pregnant and non-pregnant women, demonstrating its ability to objectively and quantitatively measure cervical stiffness^21–24^.

In this study, we assessed cervical stiffness using the Pregnolia System in largely unselected pregnant women and evaluated the predictive capabilities of CSI to assess the risk of sPTB. We refer to the population as largely unselected, since women with CL <15mm were excluded. The hypotheses of the study are that CSI is significantly lower in women delivering prematurely compared to women delivering at term, and that CSI at mid-pregnancy provides at least the same predictive power as CL.

## Material and Methods

### Clinical investigation design

This was a prospective cohort study (NCT02037334, Biomechanics Based Prediction of Preterm Delivery (SoftCervix)) conducted at 12 centers in Switzerland and one center in Belgium. The study and amendments were approved by the Cantonal Ethics Committee of Zurich (KEK-ZH-Nr. 2013-0244), Swissmedic (2013-MD-0036) and the Medical Ethics Committee of the University Hospital KU Leuven (B322201317722 (ML9440)).

The study was conducted in compliance with the protocol, the Declaration of Helsinki, the European Directive on medical devices 93/42/EEC, EN ISO14155, EN ISO14971 as well as national legal and regulatory requirements.

All study participants provided written informed consent prior to any assessment.

### Investigational device

The study was performed by using one of the following 3 device versions, and the protocol was amended whenever a new version was made available.

The first version was the Cervical Aspirator 3.1, described elsewhere^21,25^.

The second was the Cervical Aspirator 3.2 consisting of a smaller control unit and a disposable probe instead of a reusable one.

The third and last version is the Pregnolia System, the CE-marked medical device described in Badir et al^24^, functionally similar to the second version with a more user-friendly control unit.

Bench testing using silicone cervix models showed no major differences in performance among the variants.

### Study population

Women presenting at their mid-pregnancy consultation (18^0/7^ – 22^0/7^ gestational weeks) were eligible. Inclusion criteria were age (18–55 years), ability to understand nature and content of the trial and legally able to give informed consent. Exclusion criteria were communication problems, active bleeding, premature rupture of membranes (PROM), active genital infection, known carrier of HIV or Hepatitis B or C, placenta previa, Müllerian anomalies, known or suspected study non-compliance, drug or alcohol abuse, cerclage or pessary in place. At the time of measurement, women were withdrawn if they presented with CL <15 mm, or frailness / critical lesions of cervix, or active bleeding/PROM, or active genital infection, or placenta previa.

### Study objectives

The primary objective was to quantify the difference in CSI at mid-pregnancy between women with term delivery and those with sPTB. The secondary objective was to evaluate predictive capabilities of CSI measured with the Pregnolia System.

### Study procedures

After providing informed consent, CL was measured according to the Fetal Medicine Foundation protocol and the cervical consistency index (CCI) was measured as described in Parra‐Saavedra et al^20,26^. Briefly, CCI is the ratio between the manually compressed an uncompressed diameter of the cervix assessed with a transvaginal ultrasound probe.

Lastly, CSI was measured 3 consecutive times at the same location, with the probe tip slightly detached from the cervical tissue between repetitions.

Women were asked to assess the pain and discomfort of the CSI and CCI assessments with a rating from 0 (no pain or discomfort) to 10 (maximum possible pain).

The study included only one visit per woman. Birth data were collected in absence of the woman.

### Sample size estimation

Among healthy women^21^, the second trimester mean CSI was 77 mbar ± 39 mbar^21^ and according to prior data^20^ the stiffness in women with PTB was expected to be 30% lower. Power analysis calculated an effect size of 0.69. With α=0.05 and power of 0.95, at least 50 women in each group are needed to measure the difference significantly. Considering a sPTB rate of ∼5%^27,28^, we calculated that about 1000 women were needed to obtain ∼50 sPTB and ∼950 term deliveries.

### Statistical analysis

Out of the three CSI measurements, the mean value was used for the analyses. Results are reported as median (interquartile range (IQR), using R quantile function, type 7), unless otherwise specified.

CSI values are lognormally distributed. For statistical significance and p-value determination we applied the Wilcoxon Rank Sum Test when comparing two groups on a continuous variable, Fisher’s Exact Test for categorical dichotomous variables, and Pearson’s Chi-squared Test to evaluate the independence between the groups on categorical variables with more than two possible values, as appropriate. Probability density distributions were calculated based on the maximum-likelihood fitting to a lognormal distribution. A p-value of <0.05 was considered significant. All analysis was performed in R (version 4.3.2)^29^ using the R stats package for statistical significance determination and model evaluation.

Boxplots visualize the median, two hinges corresponding to the first and third quartiles and whiskers, extending from the hinge to the highest/lowest value no further than 1.5^*^IQR from the hinge.

Combined performance of CSI and CL was tested using a generalized linear model, using the glm function from R’s stats library with mean CSI and CL as independent variables.

Additional analyses of sub-populations were performed: women with singleton gestations, women with and without history of vaginal delivery, and women with and without cervical conization were analyzed separately.

## Results

From April 2014 to January 2022, 2518 pregnant women were invited to take part in the study. In January 2022, 1002 women had been included, with the last woman out of the study in June 2022. Figure 1 illustrates the women’s disposition.

**Figure 1:**
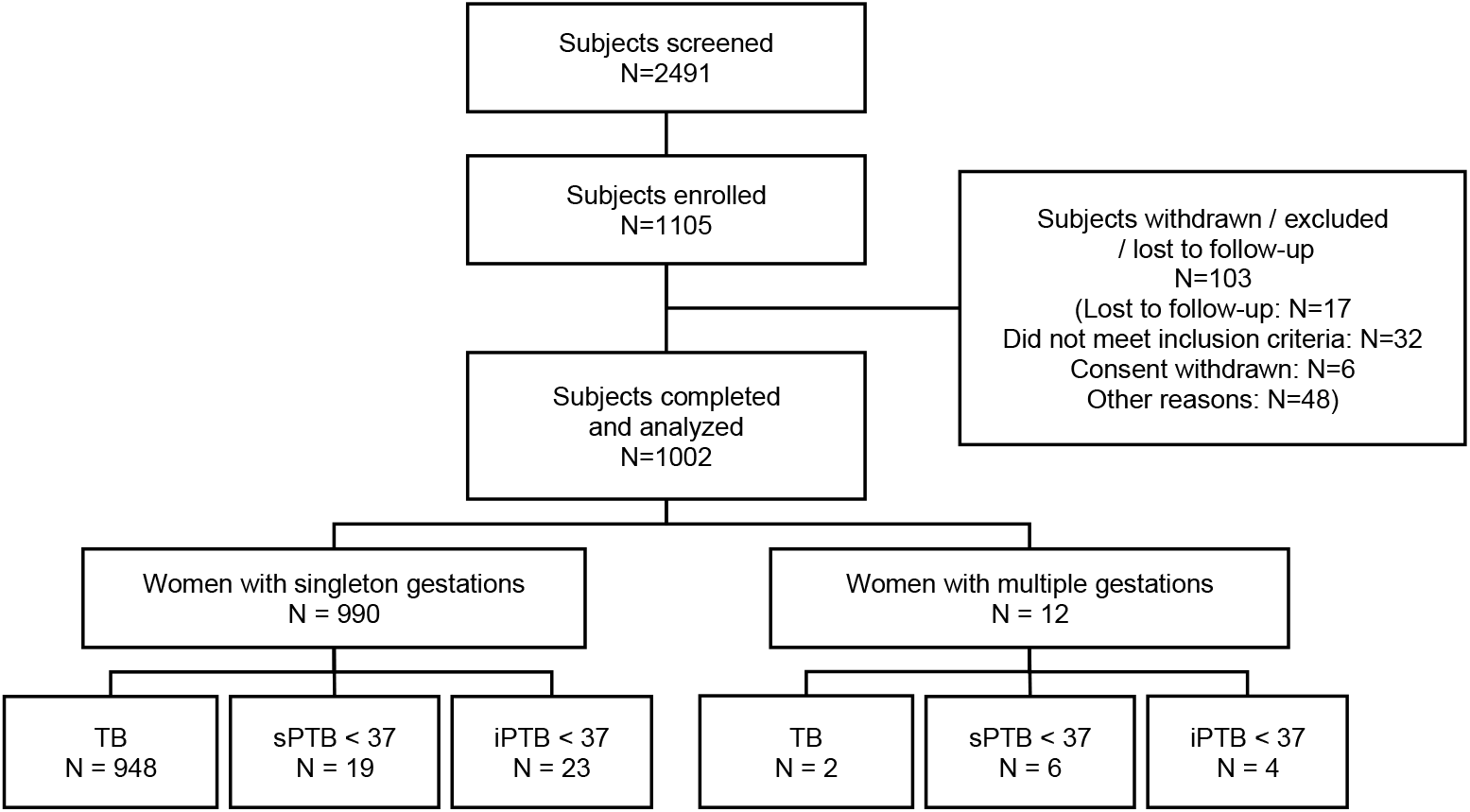
Flowchart showing participant recruitment details. TB = term birth; sPTB = spontaneous preterm birth; iPTB = iatrogenic preterm birth.

In our study, the reported PTB rate was 5.2%, with a total of 52 PTB <37 weeks, of which 48.1% (n=25) spontaneous (sPTB rate 2.5%) and 51.9% (n=27) iatrogenic (iPTB rate 2.7%). iPTB are excluded from the following analysis.

Participant demographic and clinical outcome characteristics are reported in Table 1 and Supplementary Table 1. No significant differences in any of the maternal characteristic between women delivering at term (term birth, TB) and women with a sPTB were observed.

**Table 1:** Women characteristics and birth outcomes.

| Characteristic | TB<br>N = 950<sup>1</sup> | sPTB < 37<br>weeks<br>N = 25<sup>1</sup> | p-value |
| --- | --- | --- | --- |
| Ethnicity |  |  |  |
| Caucasian | 887 (93.3%) | 21 (84.0%) |  |
| Asian | 33 (3.5%) | 1 (4.0%) |  |
| African | 19 (2.0%) | 1 (4.0%) |  |
| Other | 11 (1.2%) | 2 (8.0%) |  |
| Age at measurement (years) | 33.1 (30.3, 35.9) | 34.1 (30.6, 37.1) | 0.423<sup>2</sup> |
| Body Mass Index | 22.6 (20.6, 25.4) | 22.2 (20.1, 24.1) | 0.598<sup>2</sup> |
| Parity |  |  | 0.240<sup>3</sup> |
| 0 | 457 (48.1%) | 15 (60.0%) |  |
| ≥ 1 | 493 (51.9%) | 10 (40.0%) |  |
| Previous vaginal delivery |  |  | 0.318<sup>3</sup> |
| 0 | 591 (62.2%) | 18 (72.0%) |  |
| ≥ 1 | 359 (37.8%) | 7 (28.0%) |  |
| # Fetuses |  |  | <0.001 |
| 1 | 948 (99.8%) | 19 (76.0%) |  |
| 2 | 2 (0.2%) | 5 (20.0%) |  |
| 3 | 0 (0.0%) | 1 (4.0%) |  |
| Prior PTB |  |  | >0.999 |
| No | 903 (95.1%) | 24 (96.0%) |  |
| Yes | 47 (4.9%) | 1 (4.0%) |  |
| Prior sPTB |  |  | >0.999 |
| No | 938 (98.7%) | 25 (100.0%) |  |
| Yes | 12 (1.3%) | 0 (0.0%) |  |
| Prior conization | 17 (1.8%) | 2 (8%) | 0.083<sup>4</sup> |
| Current smoker | 51 (5.4%) | 3 (12.0%) | 0.156<sup>4</sup> |
| Alcohol intake during pregnancy | 11 (1.2%) | 1 (4.0%) | 0.269<sup>4</sup> |
| GA at measurement | 20<sup>+5</sup> (20<sup>+2</sup>, 21<sup>+2</sup>) | 20<sup>+4</sup> (20<sup>+1</sup>, 21<sup>+0</sup>) | 0.218<sup>2</sup> |
| Onset of labor |  |  | <0.001 |
| spontaneous | 600 (63.2%) | 25 (100.0%) |  |
| induced | 350 (36.8%) | 0 (0.0%) |  |
| Mode of delivery |  |  | 0.581<sup>4</sup> |
| Spontaneous vaginal delivery | 577 (60.7%) | 14 (56.0%) |  |
| Vaginal operative | 251 (26.4%) | 9 (36.0%) |  |
| C-section (primary or secondary) | 122 (12.9%) | 2 (8.0%) |  |
| GA at delivery<sup>5</sup> | 39<sup>+4</sup> (37<sup>+0</sup>, 42<sup>+1</sup>) | 35<sup>+0</sup> (32<sup>+0</sup>, 36<sup>+6</sup>) | <0.001 |
| Birth weight (g) | 3,360 (3,070, | 2,430 (2,180, | <0.001 |

<sup>1</sup>n (%); Median (IQR)
<sup>2</sup>Wilcoxon rank sum test <sup>3</sup>Pearson's Chi-squared test <sup>4</sup>Fisher's exact test
<sup>2</sup>Fisher's exact test; Wilcoxon rank sum test; Pearson's Chi-squared test
<sup>5</sup>Mean (range)
TB: term birth; sPTB: spontaneous preterm birth; PTB: preterm birth; fertilization; GA: gestational age.

There were no serious adverse events related to the aspiration device. The pain scores were 0.51±1.32 for CSI and 0.41±1.04 for CCI (mean ± standard deviation), showing that both assessments were very well tolerated with no or minimal discomfort.

The mean CSI obtained with each of the three versions was found not to be statistically different (Figure 2), confirming that the three versions are equivalent, and the data obtained can be pooled for analysis.

**Figure 2:**
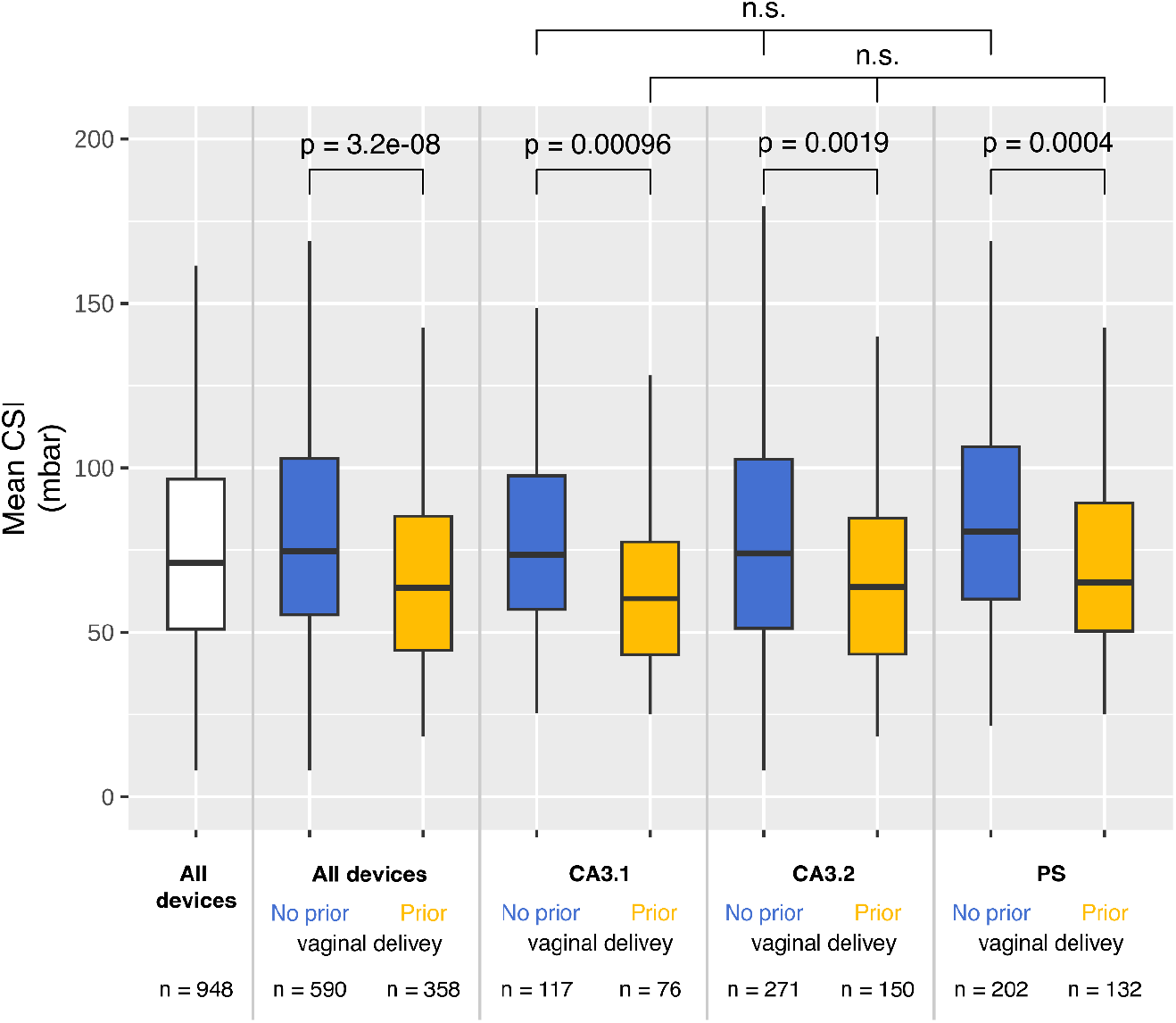
Box plots with mean CSI in women with singleton gestation. Data is reported pooled for all devices used (left), and stratified for each device version in women with and without prior vaginal delivery (All devices, Cervical Aspirator 3.1 - CA3.1, Cervical Aspirator 3.2 -CA3.2 and Pregnolia System - PS) as well. Women with previous vaginal delivery have a significantly softer cervix. The CSI distributions in women with and without prior vaginal deliveries are remarkably similar across different devices. n.s., not significant.

When considering all pregnancies (singleton and multiple), no statistically significant difference in any of the parameters was found (Table 2, left). However, analyzing singleton pregnancies only, mean CSI in women with a TB (n=948, 71 mbar (51,97)) was significantly different than mean CSI in women with a sPTB (n=19, 65 mbar (42,70)) (Table 2, right). CL and CCI do not show a significant difference between the two groups.

**Table 2:**
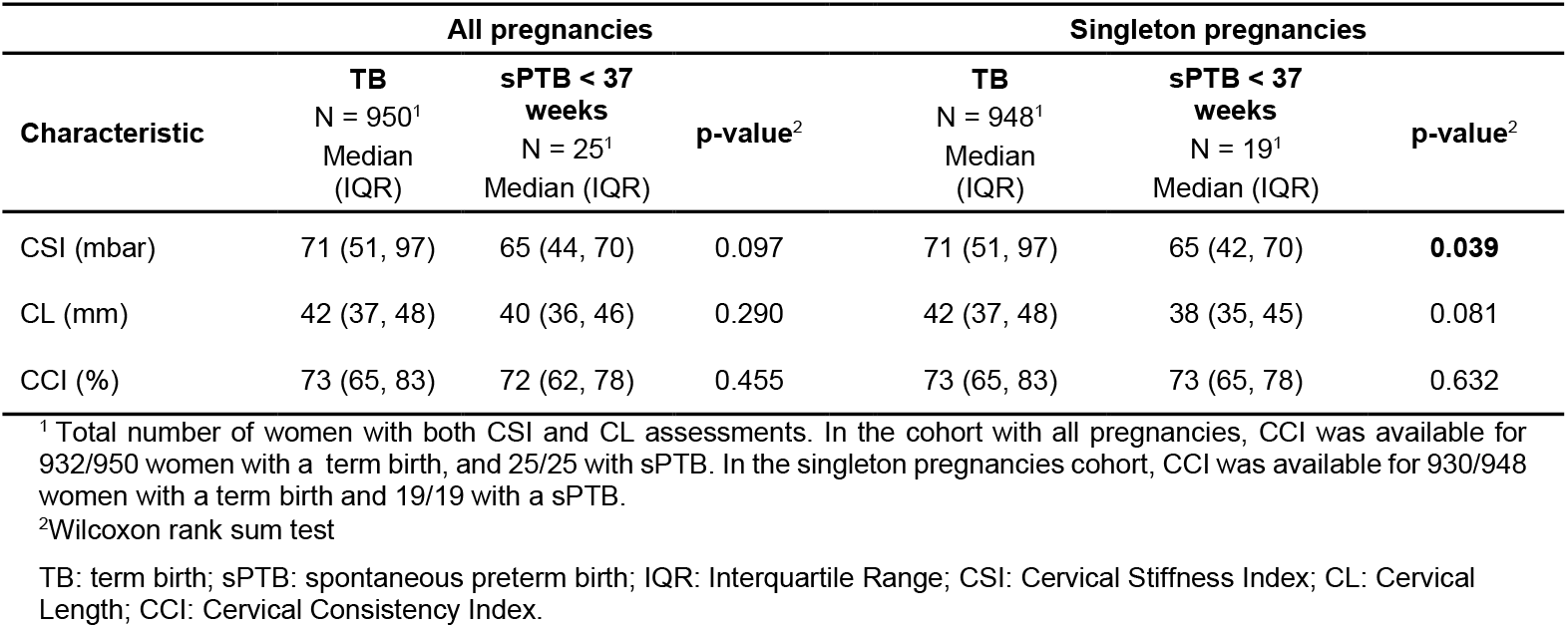
Pregnancy outcome and measurement results for singleton and multiple pregnancies and singleton pregnancies only.

| Characteristic | All pregnancies |  |  | Singleton pregnancies |  |  |
| --- | --- | --- | --- | --- | --- | --- |
|  | TB<br>N = 950<sup>1</sup><br>Median<br>(IQR) | sPTB < 37<br>weeks<br>N = 25<sup>1</sup><br>Median (IQR) | p-value<sup>2</sup> | TB<br>N = 948<sup>1</sup><br>Median<br>(IQR) | sPTB < 37<br>weeks<br>N = 19<sup>1</sup><br>Median (IQR) | p-value<sup>2</sup> |
| CSI (mbar) | 71 (51, 97) | 65 (44, 70) | 0.097 | 71 (51, 97) | 65 (42, 70) | **0.039** |
| CL (mm) | 42 (37, 48) | 40 (36, 46) | 0.290 | 42 (37, 48) | 38 (35, 45) | 0.081 |
| CCI (%) | 73 (65, 83) | 72 (62, 78) | 0.455 | 73 (65, 83) | 73 (65, 78) | 0.632 |

<sup>1</sup> Total number of women with both CSI and CL assessments. In the cohort with all pregnancies, CCI was available for 932/950 women with a term birth, and 25/25 with sPTB. In the singleton pregnancies cohort, CCI was available for 930/948 women with a term birth and 19/19 with a sPTB.
<sup>2</sup>Wilcoxon rank sum test
TB: term birth; sPTB: spontaneous preterm birth; IQR: Interquartile Range; CSI: Cervical Stiffness Index; CL: Cervical Length; CCI: Cervical Consistency Index.

Furthermore, among women with a singleton pregnancy delivering at term, CSI of women without prior vaginal delivery (VD) is significantly higher than CSI of women with history of VD (Figure 2 and Table 3). Overall, mean CSI is 15% lower in women who previously delivered vaginally, and 17%, 14%, and 15% lower for the three different groups of women measured with different device versions, demonstrating that the findings are consistent.

**Table 3:** CSI, CL, and CCI (median) in women stratified for history of vaginal delivery.

| Characteristic | TB |  |  | sPTB < 37 weeks |  |  |
| --- | --- | --- | --- | --- | --- | --- |
|  | No prior VD | Prior VD | p-value<sup>2</sup> | No prior VD | Prior VD | p-value<sup>2</sup> |
|  | N = 590<sup>1</sup><br>Median (IQR) | N = 358<sup>1</sup><br>Median (IQR) |  | N = 15<sup>1</sup><br>Median (IQR) | N = 4<sup>1</sup><br>Median (IQR) |  |
| CSI (mbar) | 75 (55, 103) | 64 (45, 85) | <0.001 | 59 (42, 70) | 67 (58, 79) | 0.582 |
| CL (mm) | 42 (37, 47) | 42 (38, 48) | 0.172 | 38 (37, 44) | 37 (29, 45) | 0.515 |
| CCI (%) | 73 (66, 83) | 72 (64, 82) | 0.124 | 67 (62, 80) | 76 (74, 78) | 0.342 |

<sup>1</sup> Total number of women with both CSI and CL assessments. CCI value was available for 578/590 women with a term birth without prior vaginal delivery and 352/358 women with a term birth with prior vaginal delivery
<sup>2</sup>Wilcoxon rank sum test
TB: term birth; sPTB: spontaneous preterm birth; IQR: Interquartile Range; VD: vaginal delivery; CSI: Cervical Stiffness Index; CL: Cervical Length; CCI: Cervical Consistency Index.

In contrast, no significant differences were found for CL or CCI.

Among women with a singleton pregnancy and a sPTB, no significant difference in CSI, CL and CCI was found between those with a prior VD (n=4) and those without history of VD (n=15). Of note, the number of women in these subgroups is low.

In the group of women with no history of VD and a singleton pregnancy, mean CSI shows a statistically significant difference between women with a TB (n=590) and women with a sPTB (n=15), as shown in Table 4. In contrast, in the group with prior VD, no statistically significant difference was found in any of the examined parameters; it is noteworthy, however, that in this sub-group only 4 women were delivering prematurely and 358 at term.

**Table 4:** Pregnancy outcome and measurements data in women with singleton pregnancy and a history of vaginal delivery.

| Characteristic | No prior VD |  |  | Prior VD |  |  |
| --- | --- | --- | --- | --- | --- | --- |
|  | TB | sPTB < 37 weeks | p-value<sup>2</sup> | TB | sPTB < 37 weeks | p-value<sup>2</sup> |
|  | N = 590<sup>1</sup><br>Median (IQR) | N = 15<sup>1</sup><br>Median (IQR) |  | N = 358<sup>1</sup><br>Median (IQR) | N = 4<sup>1</sup><br>Median (IQR) |  |
| CSI (mbar) | 75 (55, 103) | 59 (42, 70) | 0.007 | 64 (45, 85) | 67 (58, 79) | 0.872 |
| CL (mm) | 42 (37, 47) | 38 (37, 44) | 0.224 | 42 (38, 48) | 37 (29, 45) | 0.203 |
| CCI (%) | 73 (66, 83) | 67 (62, 80) | 0.324 | 72 (64, 82) | 76 (74, 78) | 0.409 |

<sup>1</sup> Total number of women with both CSI and CL assessments. CCI values were available for 578 women with a term birth and without prior vaginal delivery and 352 women with a term birth and with prior vaginal delivery.
<sup>2</sup>Wilcoxon rank sum test
VD: vaginal delivery; TB: term birth; sPTB: spontaneous preterm birth; IQR: Interquartile Range; CSI: Cervical Stiffness Index; CL: Cervical Length; CCI: Cervical Consistency Index.

Figure 3 shows CSI distributions for women in Switzerland with singleton delivery at term in our study (n=950) and in the study from Badir et al^21^ (n=42), measured at the same gestational age. The two distributions overlap considerably. Supplementary Figure 1 additionally shows CSI distributions earlier and later in pregnancy from that study, illustrating the dynamic behavior of CSI throughout gestation. We also compared three individual sets of measurements for single investigator-device version combinations totaling more than 50 measurements, which yielded that the results were not-significantly different (Supplementary Table 2).

**Figure 3:**
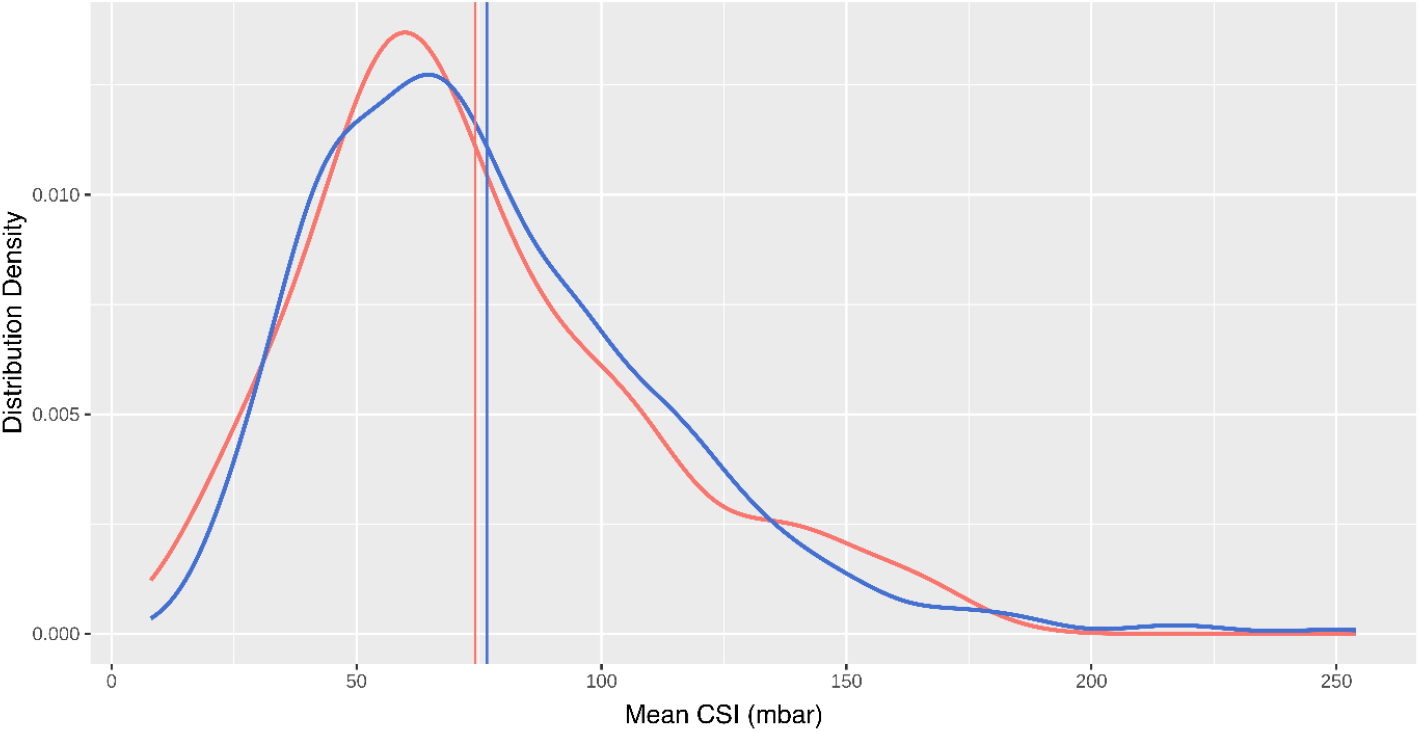
CSI log normal probability density distributions at 18^+0^ - 21^+6^ weeks for women with a singleton gestation who delivered at term. The two curves overlap considerably, indicating consistency of the results obtained in our study (blue, n = 950) and from Badir et al^21^ (red, n=42). Vertical lines indicate the distribution medians.

Figure 4 summarizes the CSI differences between women delivering at term and women spontaneously delivering before 37 weeks.

**Figure 4:**
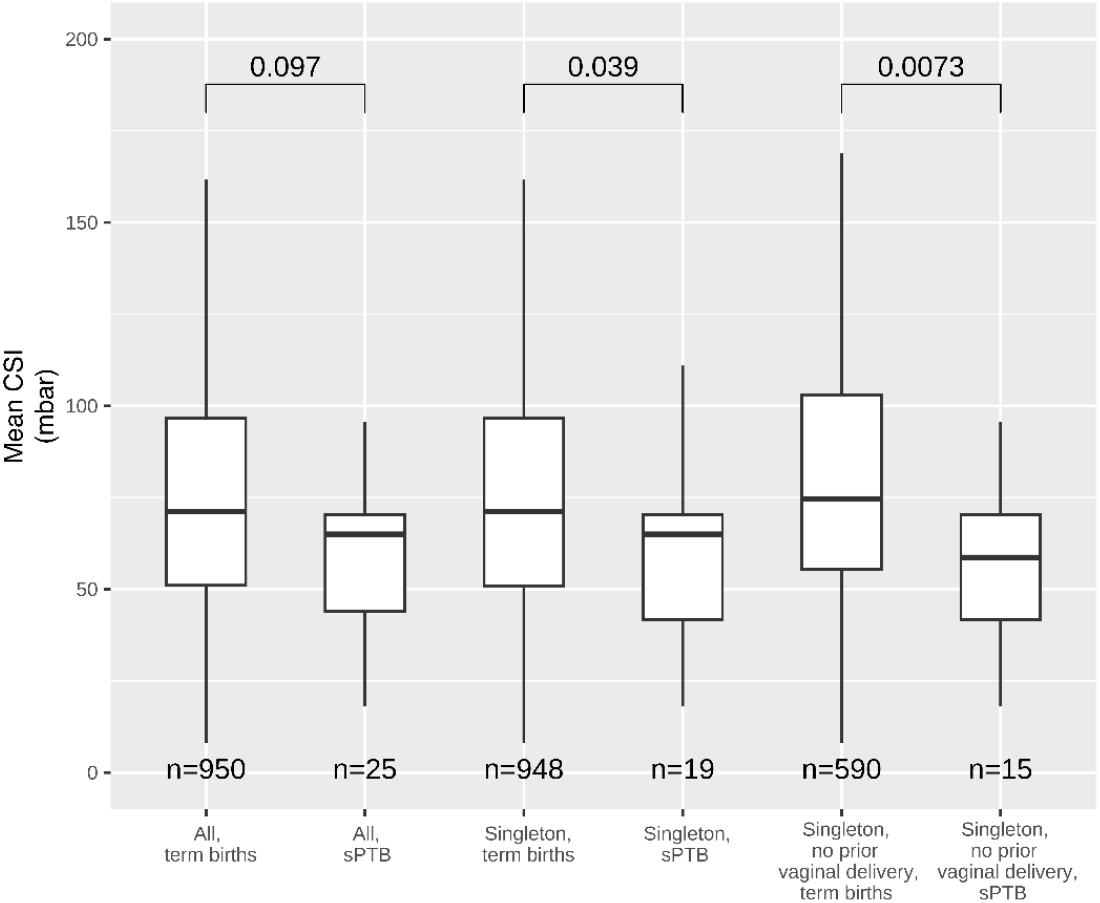
Box plots of CSI showing the differences between deliveries at term and before 37 weeks (sPTB) for singleton and multiple gestations (left), singleton gestations only (center), and singleton gestations without history of vaginal delivery (right).

In the sub-group of women with a singleton gestation and without prior VD, CSI shows a prediction improvement compared to CL (AUC 0.703 vs 0.592, respectively) (Figure 5). Combining CSI and CL in a generalized linear model provides a marginal improvement (AUC 0.729); however, CL was not considered a significant term in the model (p=0.25).

**Figure 5:**
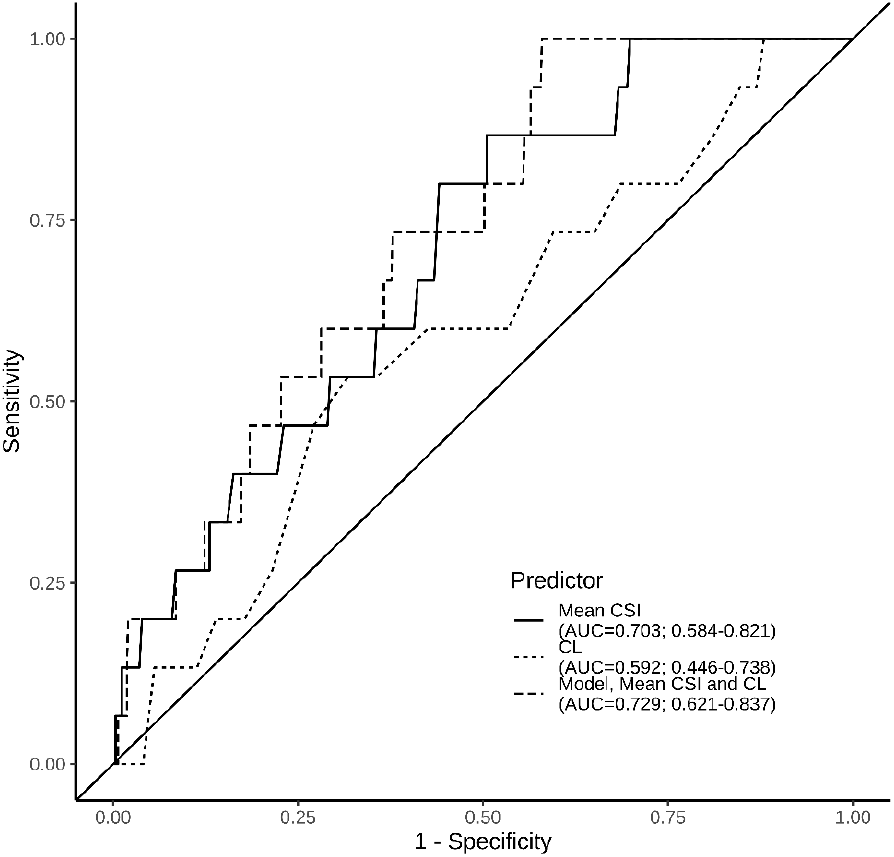
ROC curves for spontaneous preterm birth prediction in women with a singleton gestation and without a history of vaginal delivery. Areas under the ROC curves (AUC) are: mean CSI 0.703 (95% CI, 0.584 – 0.821), CL 0.592 (95% CI, 0.446 – 0.738), model including CSI and CL 0.729 (95% CI, 0.621-0.837).

The performance characteristics of different cut-offs for CSI and CL to predict sPTB in women with singleton pregnancy and no prior VD are reported in Table 5. CSI appears to be a better predictor than CL. When using the standard CL cut-off at mid-gestation (25 mm), the sensitivity of CL is 0%, indicating that none of the women at risk would be identified. However, women with CL <15 mm were excluded, therefore this comparison is valid in this CL range only. CSI cut-offs were selected to showcase sensitivity for high specificity values. Performance in women with a prior VD and a singleton gestation was not analyzed due to the low number of sPTB in this cohort (n=4).

**Table 5:** Performance of different cut-offs of CSI and CL to predict sPTB in women with singleton gestation and no prior vaginal delivery (n = 590 TB and n = 15 sPTB). Reported values and 95% confidences intervals.

| Cut-off | Sensitivity | Specificity | PPV | NPV | TP | FP | TN | FN | Prevalence |
| --- | --- | --- | --- | --- | --- | --- | --- | --- | --- |
| **Mean CSI (mbar)** |  |  |  |  |  |  |  |  |  |
| 30 | 20% (0, 40.2) | 96.1% (94.5, 97.7) | 11.5% (0, 23.8) | 97.9% (96.8, 99.1) | 3 | 23 | 567 | 12 | 4.3% |
| 40 | 26.7% (4.3, 49.0) | 91.2% (88.9, 93.5) | 7.14% (0.4, 13.9) | 98% (96.8, 99.2) | 4 | 52 | 538 | 11 | 9.3% |
| 50 | 40% (15.2, 64.8) | 80.3% (77.1, 83.5) | 4.92% (1.1, 8.8) | 98.1% (96.9, 99.3) | 6 | 116 | 474 | 9 | 20.2% |
| **CL (mm)** |  |  |  |  |  |  |  |  |  |
| 25 | 0% (0.0, 0.0) | 99.7% (99.2, 100) | 0% (0.0, 0.0) | 97.5% (96.3, 98.8) | 0 | 2 | 588 | 15 | 0.3% |
| 30 | 0% (0.0, 0.0) | 97.6% (96.4, 98.9) | 0% (0.0, 0.0) | 97.5% (96.2, 98.7) | 0 | 14 | 576 | 15 | 2.3% |
| 35 | 20% (0.0, 40.2) | 86.1% (83.3, 88.9) | 3.5% (0.0, 7.45) | 97.7% (96.4, 99.0) | 3 | 82 | 508 | 12 | 14.0% |

PPV: positive predictive value, NPV: negative predictive value, TP: true positives, FP: false positives, TN: true negatives, FN: false negatives; CSI: Cervical Stiffness Index; CL: Cervical Length. Prevalence: percentage of women with a soft or short cervix for each threshold.

Table 6 shows that women with a singleton pregnancy delivering at term and with prior conization have a statistically significantly stiffer and a (not significantly) shorter cervix than women without prior conization.

**Table 6:**
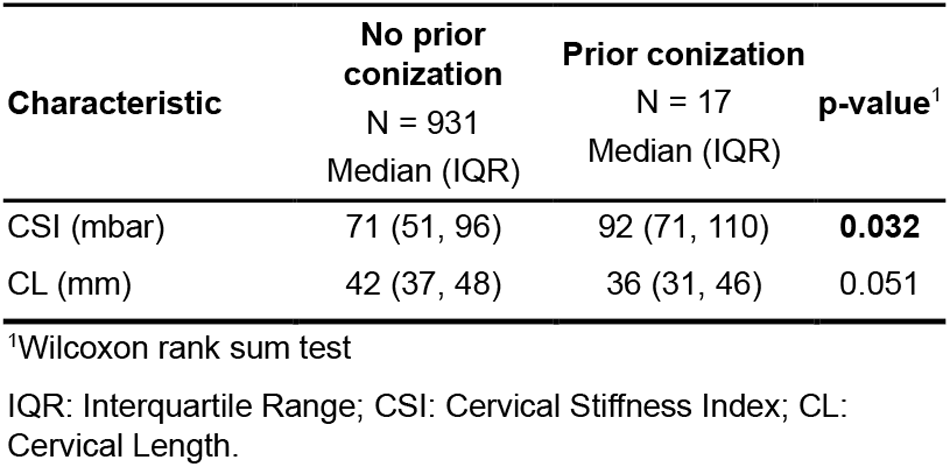
CSI difference in women with a singleton pregnancy and a TB without and with prior conization.

| Characteristic | No prior conization<br>N = 931<br>Median (IQR) | Prior conization<br>N = 17<br>Median (IQR) | p-value<sup>1</sup> |
| --- | --- | --- | --- |
| CSI (mbar) | 71 (51, 96) | 92 (71, 110) | **0.032** |
| CL (mm) | 42 (37, 48) | 36 (31, 46) | 0.051 |

<sup>1</sup>Wilcoxon rank sum test
IQR: Interquartile Range; CSI: Cervical Stiffness Index; CL: Cervical Length.

Lastly, we found no influence of treatment for PTB prevention on CSI (Supplementary Table 3).

## Discussion

### Principal findings

In this study, the cervical stiffness of 1002 largely unselected pregnant women presenting at their mid-pregnancy consultation was measured with an aspiration-based device and was associated with gestational age at birth. Measurements with the Pregnolia System were painless and safe. The reported PTB rate of 5.2% was lower than the Swiss national rate (6.4% in 2020^30^). Nevertheless, in singleton pregnancies, CSI was significantly softer in women with sPTB compared to women with a TB and appears to be a better predictor of sPTB compared to CL (Supplementary Figure 2). Besides, a significant and robust decrease in CSI was associated with history of VD, compared to nullipara or women who delivered through cesarian section. This suggests a mechanism: prior VD alters the mechanical properties of the cervix, leading to a softer structural constitution of the organ. Furthermore, device performance was considered robust, delivering consistent results among subgroups of different women measured with different devices and by different investigators, as well as with a previous study.

### Results in the context of what is known

At mid-gestation, women with a singleton gestation delivering at term have a significantly stiffer cervix than women with a sPTB. This finding is in line with the literature, whilst cervical stiffness is measured with other methods, for example using E-Cervix^10,11^, shear wave elastography (SWE)^14,15^, or the CCI method^18,20^.

CSI in women without a history of VD was found to be significantly higher than in women with prior VD. To the best of our knowledge, this is the first quantification of the reduction in cervical stiffness due to prior VD as other studies have only assessed differences in stiffness between nulliparous and multiparous women. Duan et al.^31^ reported that the stiffness measured with SWE at the external os of nulliparous women is significantly higher than the stiffness of multiparous women without previous PTB, while Zhang et al.^32^ did not see any difference in SWE parameters of both internal cervical os and external cervical os among nulliparous, primiparous and multiparous women.

In our study, among women with a singleton pregnancy, the decrease in CSI between the ones with a TB and those with a sPTB was 8% and 15% in the sub-group of women without prior VD. Other authors reported a decrease in stiffness in women delivering prematurely between 7% and 30%^10,11,18,20^. Notably, ultrasound-based techniques measure the cervical stiffness at the internal os, middle cervix and external os. In contrast, our technique measures the stiffness at the vaginally exposed portion of the cervix, namely at the anterior lip. Our results demonstrate the measurement can be conducted locally.

A comparison of the performance of CSI in this study with performance of stiffness assessed with other technologies, in studies in unselected women or with a low-risk pregnancy of similar size (at least 500 women or ≥30 sPTB) was conducted (Supplementary Table 4). PTB predictive performance is overall similar for similar trials, although smaller trials with higher sPTB rate^33,34^ or case-controlled^35,36^ ones show noticeable higher predictive performance. It should be noticed, however, that most of these trials are monocenter.

A large discrepancy was seen between published CCI results^18,20,34^ and those obtained within this trial. It is not immediately clear why; however, it could be due to training levels and a higher number of clinicians measuring.. This may indicate that measuring stiffness with the Pregnolia System is easier and requires less specialized training than with the CCI method.

In this study, where women with CL <15 mm were excluded, the AUC of CL for prediction of sPTB in singleton pregnancies without prior VD was 0.592. When comparing the AUC for CL in this study with AUC for CL in studies including unselected or low-risk women with singleton gestations, where very short CL were not excluded, they are found to be similar (Supplementary Table 5).

### Clinical implications

The cervix plays a fundamental role in ensuring that a pregnancy is carried out to term. When it is no longer structurally sound (e.g. abnormal shortening, cervical insufficiency), it cannot perform its function adequately, potentially leading to premature labor^37^. Multiple etiologies can lead to such failure, but the overall knowledge of what leads to cervical failure is far from complete. With this study, we have addressed cervical stiffness and how it correlates with sPTB in largely unselected pregnant women, reinforcing the body of evidence that at least some of them have an abnormally soft cervix and do not necessarily have a shortened cervix.

### Research implications

In this study, the combination of CSI and other parameters in multivariate models has not improved the predictive capability of CSI alone. Nevertheless, our results strengthen the potential for considering quantifiable cervical stiffness as a relevant parameter for cervical status assessment and evaluation of PTB risk. However, due to the multifactorial nature of PTB, the combination of CSI with other risk factors shall be further and systematically investigated. Moreover, the assessment of CSI in other cohorts, such as high-risk pregnancies or symptomatic women shall be evaluated.

### Strength and findings

This study has several strengths. First, contrary to studies with only one or a few investigators (Supplementary Table 4), CSI was measured in 13 different centers by more than 45 investigators (Supplementary Table 6), providing robustness to the findings and the confidence that these results can be easily replicated by others. Second, this was a large, prospective study assessing cervical stiffness in a largely unselected general population. By analyzing the characteristics of the those delivering at term, we determined that baseline CSI is affected by prior VD. This finding is consistent with the overall impression of clinicians who palpate the cervix and provides a robust determination of the effect of the first VD on cervical stiffness.

Our study also has limitations. First, the study was underpowered, as we only observed half of the expected sPTBs. Second, the possibility of performing sub-group analysis was limited by the low number of cases in each sub-group.

### Conclusion

To our knowledge, this is the first study assessing cervical stiffness at mid-gestation with an aspiration-based device in largely unselected women and correlating it with the risk of delivering prematurely. Furthermore, we quantified a robust softening of cervical tissue in women with a history of VD. Cervical stiffness measurement with the Pregnolia System is safe and painless and has shown promise as a predictor for sPTB in this cohort.

## Data Availability

Author elects to not share data.

## Acknowledgment

The authors would like to thank all the women who took part in this study and the collaborators who made the recruitment possible, as well as Dr. Laura Bernardi (Pregnolia AG) for reviewing, and Dr. Michael Fernandez (Pregnolia AG), Dr. Francisco Delgado (Pregnolia AG), and Dr. Erik P. Willems for the statistical analysis.

## Disclosure

S.B. declares competing financial interests as founder of Pregnolia AG. G.H. and K.Q.L. report research grants for other clinical investigations with the Pregnolia System. This research is funded, in part, by a grant from Pregnolia AG. Pregnolia AG provided devices to all participating sites. The remaining authors report no conflict of interest. D.S. is the sponsor investigator.

## Funding

The sponsor has received internal funding of Federal Institute of Technology Zurich, a grant from the Swiss National Science Foundation and a grant from Pregnolia AG.

Clinical trial registration: NCT02037334 https://clinicaltrials.gov/study/NCT02037334

## Notes

### Clinical Trial

NCT02037334

### Author Declarations

The study and amendments were approved by the Cantonal Ethics Committee of Zurich (KEK-ZH-Nr. 2013-0244), Swissmedic (2013-MD-0036) and the Medical Ethics Committee of the University Hospital KU Leuven (B322201317722 (ML9440)).

## References

1. World Health Organization. Born Too Soon: Decade of Action on Preterm Birth. (WHO, UNFPA, UNICEF P, ed.).; 2023. https://www.who.int/publications/i/item/9789240073890

2. Vidal MS, Lintao RCV, Severino MEL, Tantengco OAG, Menon R. Spontaneous preterm birth: Involvement of multiple feto-maternal tissues and organ systems, differing mechanisms, and pathways. Front Endocrinol (Lausanne).Frontiers Media S.A. 2022;13. doi:10.3389/fendo.2022.1015622

3. Breslin N, Gyamfi-Bannerman C. Current Preterm Birth Prevention Strategies. Clin Perinatol.W.B. Saunders. 2020;47(4):705–717. doi:10.1016/j.clp.2020.08.001

4. Van Der Ven J, Van Os MA, Kazemier BM, et al. The capacity of mid-pregnancy cervical length to predict preterm birth in low-risk women: A national cohort study. Acta Obstet Gynecol Scand. 2015;94(11):1223–1234. doi:10.1111/aogs.12721

5. Iams JD, Goldenberg RL, Mercer BM, et al. The preterm prediction study: can low-risk women destined for spontaneous preterm birth be identified? Am J Obstet Gynecol. 2001;184(4):652–655. doi:10.1067/mob.2001.111248

6. Costantine MM, Ugwu L, Grobman WA, et al. Cervical length distribution and other sonographic ancillary findings of singleton nulliparous patients at midgestation. Am J Obstet Gynecol. 2021;225(2):181–e1. doi:10.1016/j.ajog.2021.02.017

7. Mazza E, Parra-Saavedra M, Bajka M, Gratacos E, Nicolaides K, Deprest J. In vivo assessment of the biomechanical properties of the uterine cervix in pregnancy. Prenat Diagn. 2014;34(1):33–41. doi:10.1002/pd.4260

8. Feltovich H. Cervical Evaluation: From Ancient Medicine to Precision Medicine. Obstetrics and Gynecology. 2017;130(1):51–63. doi:10.1097/AOG.0000000000002106

9. Vink J, Feltovich H. Cervical etiology of spontaneous preterm birth. Semin Fetal Neonatal Med. 2016;21(2):106–112. doi:10.1016/j.siny.2015.12.009

10. Du L, Zhang LH, Zheng Q, et al. Evaluation of Cervical Elastography for Prediction of Spontaneous Preterm Birth in Low-Risk Women: A Prospective Study. Journal of Ultrasound in Medicine. 2020;39(4):705–713. doi:10.1002/jum.15149

11. Liu Y, Yang D, Jiang Y, Yue Q. Quantification of cervical stiffness changes in single and twin pregnancies using the E-Cervix technique. Am J Obstet Gynecol MFM. 2023;5(2):100804. doi:10.1016/j.ajogmf.2022.100804

12. Patberg ET, Wells M, Vahanian SA, et al. Use of cervical elastography at 18 to 22 weeks’ gestation in the prediction of spontaneous preterm birth. Am J Obstet Gynecol. 2021;225(5):525.e1-525.e9. doi:10.1016/j.ajog.2021.05.017

13. Hernandez-Andrade E, Aurioles-Garibay A, Garcia M, et al. Effect of depth on shear-wave elastography estimated in the internal and external cervical os during pregnancy. J Perinat Med. 2014;42(5):549–557. doi:10.1515/jpm-2014-0073

14. Hernandez-Andrade E, Maymon E, Luewan S, et al. A soft cervix, categorized by shear-wave elastography, in women with short or with normal cervical length at 18-24 weeks is associated with a higher prevalence of spontaneous preterm delivery. J Perinat Med. 2018;46(5):489–501. doi:10.1515/jpm-2018-0062

15. O’Hara S, Zelesco M, Sun Z. Use of shear wave elastography on the maternal cervix to recognise cervical insufficiency using a transabdominal ultrasound approach. Australas J Ultrasound Med. 2021;24(2):89–98. doi:10.1002/ajum.12236

16. Yang X, Ding Y, Mei J, et al. Second-Trimester Cervical Shear Wave Elastography Combined With Cervical Length for the Prediction of Spontaneous Preterm Birth. Ultrasound Med Biol. 2022;48(5):820–829. doi:10.1016/j.ultrasmedbio.2022.01.003

17. Feng Q, Chaemsaithong P, Duan H, et al. Screening for spontaneous preterm birth by cervical length and shear-wave elastography in the first trimester of pregnancy. Am J Obstet Gynecol. 2022;227(3):500.e1-500.e14. doi:10.1016/j.ajog.2022.04.014

18. Baños N, Julià C, Lorente N, et al. Mid-trimester cervical consistency index and cervical length to predict spontaneous preterm birth in a high-risk population. American Journal of Perinatology Reports. 2018;8(01):e43–e50. doi:10.1055/s-0038-1636993

19. Baños N, Murillo-Bravo C, Julià C, et al. Mid-trimester sonographic cervical consistency index to predict spontaneous preterm birth in a low-risk population. Ultrasound in Obstetrics and Gynecology. 2018;51(5):629–636. doi:10.1002/uog.17482

20. Parra-Saavedra M, Gómez L, Barrero A, Parra G, Vergara F, Navarro E. Prediction of preterm birth using the cervical consistency index. Ultrasound in Obstetrics and Gynecology. 2011;38(1):44–51. doi:10.1002/uog.9010

21. Badir S, Mazza E, Zimmermann R, Bajka M. Cervical softening occurs early in pregnancy: characterization of cervical stiffness in 100 healthy women using the aspiration technique. Prenat Diagn. 2013;33(8):737–741. doi:10.1002/pd.4116

22. Badir S, Mazza E, Bajka M. Objective Assessment of Cervical Stiffness after Administration of Misoprostol for Intrauterine Contraceptive Insertion. Ultrasound Int Open. 2016;2(2):E63–E67. doi:10.1055/s-0042-106393

23. Stone J, House M. Measurement of cervical softness before cerclage placement with an aspiration-based device. Am J Obstet Gynecol MFM. 2023;5(4):1–7. doi:10.1016/j.ajogmf.2023.100881

24. Badir S, Bernardi L, Feijó Delgado F, Quack Loetscher K, Hebisch G, Hoesli I. Aspiration technique-based device is more reliable in cervical stiffness assessment than digital palpation. BMC Pregnancy Childbirth. 2020;20(1):391. doi:10.1186/s12884-020-03080-x

25. Kyvernitakis I, Lauer P, Malan M, Badir S, Maul H. A novel aspiration technique to assess cervical remodelling in patients with or without cervical shortening: Sequence of first changes, definition of cut-off values and impact of cervical pessary, stratified for cervical length. PLoS One. 2023;18(4 April). doi:10.1371/journal.pone.0283944

26. Parra-Saavedra MA, Gómez LA, Barrero A, et al. Cervical Consistency Index: A new concept in Uterine Cervix evaluation. Donald School Journal of Ultrasound in Obstetrics and Gynecology. 2011;5(4):411–415. doi:10.5005/jp-journals-10009-1218

27. Walani SR. Global burden of preterm birth. International Journal of Gynecology and Obstetrics. 2020;150(1):31–33. doi:10.1002/ijgo.13195

28. Menon R. Spontaneous preterm birth, a clinical dilemma: Etiologic, pathophysiologic and genetic heterogeneities and racial disparity. Acta Obstet Gynecol Scand. 2008;87(6):590–600. doi:10.1080/00016340802005126

29. R Core Team (2024). R: A Language and Environment for Statistical Computing. R Foundation for Statistical Computing. Vienna, Austria. 2024;https://www.R-project.org. https://www.R-project.org/

30. Berger R, Abele H, Bahlmann F, et al. Prevention and Therapy of Preterm Birth. Guideline of the DGGG, OEGGG and SGGG (S2k-Level, AWMF Registry Number 015/025, September 2022) – Part 1 with Recommendations on the Epidemiology, Etiology, Prediction, Primary and Secondary Prevention of Preterm B. Geburtshilfe Frauenheilkd. 2023;83(5):547–568. doi:10.1055/a-2044-0203

31. Duan H, Chaemsaithong P, Ju X, et al. Shear-wave sonoelastographic assessment of cervix in pregnancy. Acta Obstet Gynecol Scand. 2020;99(11):1458–1468. doi:10.1111/aogs.13874

32. Zhang HP, Wu JJ, Zhang WY, Tao JZ, Ma C Bin, Zhou YQ. Evaluation of the stiffness of normal cervix and its change with different factors using transvaginal two-dimensional shear wave elastography under strict quality control. BMC Med Imaging. 2023;23(1):1–9. doi:10.1186/s12880-023-01020-7

33. Sowmiya R, Samal SK, Rathod S. Study of Cervical Consistency Index and Cervical Length during Mid-trimester for the Prediction of Spontaneous Preterm Labor in Low-risk Pregnancies. Journal of SAFOG. 2023;15(5):513–516. doi:10.5005/jp-journals-10006-2141

34. Oturina V, Hammer K, Möllers M, et al. Assessment of cervical elastography strain pattern and its association with preterm birth. J Perinat Med.Walter de Gruyter GmbH. 2017;45(8):925–932. doi:10.1515/jpm-2016-0375

35. Gesthuysen A, Hammer K, Möllers M, et al. Evaluation of cervical elastography strain pattern to predict preterm birth. Ultraschall in der Medizin. 2020;41(4):397–403. doi:10.1055/a-0865-1711

36. House M, Socrate S. The cervix as a biomechanical structure. Ultrasound in Obstetrics and Gynecology. 2006;28(6):745–749. doi:10.1002/uog.3850

37. Jiang L, Peng L, Rong M, et al. Nomogram Incorporating Multimodal Transvaginal Ultrasound Assessment at 20 to 24 Weeks’ Gestation for Predicting Spontaneous Preterm Delivery in Low-Risk Women. Int J Womens Health. 2022;14(February):323–331. doi:10.2147/IJWH.S356167

