## Supplementary material for "Cervical Stiffness in Mid-Pregnancy: A Predictive Tool Using a Novel Aspiration Device for Spontaneous Preterm Birth Risk Assessment"

#### Supplementary Tables

***Supplementary Table 1****: Full women characteristics and birth outcomes, including iatrogenic PTB.*

| Characteristic | Overall<br>N = 1,002[1] | TB<br>N = 950[1] | sPTB < 37 weeks<br>N = 25[1] | iPTB < 37 weeks<br>N = 27[1] | p-value[2] |
|---|---|---|---|---|---|
| Ethnicity | | | | | |
| Caucasian | 934 (93%) | 887 (93%) | 21 (84%) | 26 (96%) | |
| Asian | 34 (3.4%) | 33 (3.5%) | 1 (4.0%) | 0 (0%) | |
| African | 20 (2.0%) | 19 (2.0%) | 1 (4.0%) | 0 (0%) | |
| Other | 14 (1.4%) | 11 (1.2%) | 2 (8.0%) | 1 (3.7%) | |
| Age at Measurement | 33.1 (30.3, 36.0) | 33.1 (30.3, 35.9) | 34.1 (30.6, 37.1) | 33.9 (31.5, 37.5) | 0.460 |
| Body Mass Index | 22.6 (20.6, 25.4) | 22.6 (20.6, 25.4) | 22.2 (20.1, 24.1) | 23.7 (21.4, 27.1) | 0.420 |
| Total Parity | | | | | |
| 0 | 485 (48%) | 457 (48%) | 15 (60%) | 13 (48%) | |
| > 1 | 517 (52%) | 493 (52%) | 10 (40%) | 14 (52%) | |
| Vaginal Parity | | | | | 0.087 |
| 0 | 629 (63%) | 591 (62%) | 18 (72%) | 20 (74%) | |
| > 1 | 373 (37%) | 359 (38%) | 7 (28%) | 7 (26%) | |
| # Fetuses in Utero | | | | | <0.001 |
| 1 | 990 (99%) | 948 (100%) | 19 (76%) | 23 (85%) | |
| 2 | 11 (1.1%) | 2 (0.2%) | 5 (20%) | 4 (15%) | |
| 3 | 1 (<0.1%) | 0 (0%) | 1 (4.0%) | 0 (0%) | |
| Prior sPTB | | | | | 0.502 |
| History of PTB | 13 (1.3%) | 12 (1.3%) | 0 (0%) | 1 (3.7%) | |
| No history of PTB | 989 (99%) | 938 (99%) | 25 (100%) | 26 (96%) | |
| Prior Spontaneous Abortion/Stillbirth | 7 (0.7%) | 6 (0.6%) | 1 (4.0%) | 0 (0%) | 0.174 |
| Prior Conizations | 20 (2.0%) | 17 (1.8%) | 2 (8.0%) | 1 (3.7%) | 0.055 |
| Current Smoker | 55 (5.5%) | 51 (5.4%) | 3 (12%) | 1 (3.7%) | 0.263 |
| Currently Drinking Alcohol | 13 (1.3%) | 11 (1.2%) | 1 (4.0%) | 1 (3.7%) | 0.143 |
| Mode of Conception | | | | | 0.029 |
| Spontaneous | 944 (94%) | 899 (95%) | 23 (92%) | 22 (81%) | |
| Hormonal Support | 8 (0.8%) | 8 (0.8%) | 0 (0%) | 0 (0%) | |
| IUI | 9 (0.9%) | 8 (0.8%) | 1 (4.0%) | 0 (0%) | |
| IVF/ICSI | 41 (4.1%) | 35 (3.7%) | 1 (4.0%) | 5 (19%) | |

| Characteristic | Overall<br>N = 1,002[1] | TB<br>N = 950[1] | sPTB < 37 weeks<br>N = 25[1] | iPTB < 37 weeks<br>N = 27[1] | p-value[2] |
|---|---|---|---|---|---|
| GA at Measurement (weeks$^{+days}$) | $20^{+5}$ ($20^{+2}$, $21^{+2}$) | $20^{+5}$ ($20^{+2}$, $21^{+2}$) | $20^{+4}$ ($20^{+1}$, $21^{+0}$) | $20^{+5}$ ($20^{+2}$, $21^{+2}$) | 0.443 |
| Cervical Length (mm) | 42 (37, 47) | 42 (37, 48) | 40 (36, 46) | 41 (36, 45) | 0.228 |
| Onset of labor | | | | | <0.001 |
| spontaneous | 625 (62%) | 600 (63%) | 25 (100%) | 0 (0%) | |
| induced | 377 (38%) | 350 (37%) | 0 (0%) | 27 (100%) | |
| Mode of Delivery | | | | | <0.001 |
| Spontaneous vaginal delivery | 595 (59%) | 577 (61%) | 14 (56%) | 4 (15%) | |
| Vaginal operative | 282 (28%) | 251 (26%) | 9 (36%) | 22 (81%) | |
| C-section (primary or secondary) | 125 (12%) | 122 (13%) | 2 (8.0%) | 1 (3.7%) | |
| GA at Delivery (weeks$^{+days}$) | $39^{+4}$ ($38^{+5}$, $40^{+3}$) | $39^{+5}$ ($38^{+6}$, $40^{+3}$) | $35^{+5}$ ($33^{+5}$, $36^{+0}$) | $35^{+3}$ ($30^{+5}$, $36^{+3}$) | <0.001 |
| Birth weight (g) | 3,320 (3,016, 3,640) | 3,360 (3,070, 3,660) | 2,430 (2,180, 2,610) | 2,190 (1,650, 2,455) | <0.001 |

[1]n (%); Median (IQR)

[2]Fisher's exact test; Kruskal-Wallis rank sum test; Pearson's Chi-squared test

TB: term birth; sPTB: spontaneous preterm birth; iPTB: iatrogenic preterm birth; PTB: preterm birth; IUI: intrauterine insemination; IVF: in vitro fertilization; ICSI: Intracytoplasmic sperm injection; GA: gestational age.

***Supplementary Table 2:*** *Characteristics of three single investigator-device combinations, where more than 50 measurements occurred (different women). Mean/median are slightly lower for Cervical Aspirator 3.1 (CA3.1), and comparable between Cervical Aspirator 3.2 (CA3.2) and Pregnolia System device versions, but differences are not considered significant. Standard deviations are very similar. This reinforces the conclusion that there are no major differences between devices.*

| Characteristic | Device CA3.1 | Device CA3.2 | Device Pregnolia System | p-value[1] |
|---|---|---|---|---|
| N | 100 | 96 | 70 | |
| CSI Median (IQR) | 67 (46, 85) | 72 (50, 103) | 75 (57, 98) | <0.001 |
| CSI Mean (SD) | 71 (34) | 78 (37) | 81 (33) | |
| Range | 25–220 | 8–172 | 25–186 | |

[1]Kruskal-Wallis rank sum test

CSI: Cervical Stiffness Index; IQR: interquartile range: SD: standard deviation.

***Supplementary Table 3:*** *Difference in analyzed parameters when data are analyzed for presence or absence of treatment (cerclage and/or progesterone).*

| Characteristic | No cerclage or progesterone<br>N = 936[1]<br>Median (IQR) | Treatment with cerclage and/or progesterone<br>N = 31[1]<br>Median (IQR) | p-value[2] |
|---|---|---|---|
| CSI (mbar) | 71 (51, 96) | 68 (44, 88) | 0.475 |
| CL (mm) | 42 (37, 48) | 38 (32, 44) | **0.006** |
| CCI (%) | 73 (65, 83) | 73 (67, 85) | 0.287 |

[1] Total number of women with both CSI and CL assessments. For the CCI measurement, data were available for 918/936 women who did not receive treatment and 31/31 women who did.

[2]Fisher's exact test; Wilcoxon rank sum test

IQR: interquartile range; CSI: Cervical Stiffness Index; CL: Cervical Length; CCI: Cervical Consistency Index.

***Supplementary Table 4****: Comparison of performance characteristics of cervical stiffness to predict sPTB < 37 weeks with different methods. Studies including at least 500 women or ≥ 30 sPTB are considered.*

| Reference | Technology | Population | GA at meas. | Mono / multi | N | N sPTB | AUC | SN | SP |
|---|---|---|---|---|---|---|---|---|---|
| Current study | Aspiration (cut-off 40 mbar) | Singleton, no prior vaginal delivery | $18^{+0}$ - $22^{+0}$ | Multicenter (13 sites) | 590 (no prior vaginal delivery) | 15 (2.5%) | 0.703 | 26.7% | 91.2% |
| Patberg et al [12] | E-Cervix | Singleton | $18^{+0}$ - $22^{+0}$ | Monocenter | 798 | 49 (6.1%) | 0.511 – 0.573 depending on parameter | n/a | n/a |
| Jiang et al [37] | Strain elastography | Singleton | $20^{+0}$ - $24^{+0}$ | Multicenter (2 sites) | 1260 | 103 (8.2%) | For monogram including several parameters: 0.839 For stiffness only: n/a | n/a | n/a |
| Oturina et al[34] | Strain elastography | Singleton | $13^{+0}$ - $35^{+0}$ | Monocenter, case-controlled | 60 | 30 | 0.8322 | 73% (stiffness and length combined) | 93% (stiffness and length combined) |
| Gesthuysen et al[35] | Strain elastography | Singleton | $20^{+0}$ - $34^{+0}$ | Monocenter, case-controlled | 335 | 50 | 0.8059 | 52% (stiffness and length combined) | 96% (stiffness and length combined) |
| Wozniak et al[38] | Strain elastography | Singleton | $18^{+0}$ - $21^{+6}$ | Monocenter | 333 | 35 (10.5%) | n/a | 85.7% | 97.6% |
| Yang et al[16] | SWE | Singleton | $18^{+0}$ - $24^{+0}$ | Monocenter | 773 | 60 (7.8%) | 0.72 – 0.93 depending on region | n/a | n/a |
| Feng et al[17] | SWE | Singleton | $11^{+0}$ - $13^{+6}$ | Monocenter | 1035 | 54 (5.2%) | n/a | 20.4% | 91% |
| Hernandez-Andrade et al[14] | SWE | Singleton | $18^{+0}$ - $24^{+0}$ | Monocenter | 628 | 31 (4.9%) | n/a | 19.4% (stiffness and length combined) | 98.4% (stiffness and length combined) |
| Baños et al[18] | CCI (cut-off 64.6%) | Singleton | $19^{+0}$ - $24^{+6}$ | Monocenter | 532 | 22 (4.1%) | 0.84 | 77.3% | 82.7% |
| Parra-Saavedra et al[20] | CCI (at 10% screen-positive rate) | Singleton | Various GA | Monocenter | 1031 | 80 (7.8%) | 0.91 | 78.8% | 94.9% |
| Sowmiya et al[33] | CCI (cut-off 65%) | Singleton | $19^{+0}$ - $24^{+6}$ | Monocenter | 307 | 32 (10.4%) | 0.87 | 78.1% | 80.4% |

GA: gestational age; NP: nulliparous women; MP: multiparous women; AUC: area under the ROC curve; SN: sensitivity; SP: specificity; SWE: shear wave elastography; CCI: cervical consistency index.

***Supplementary Table 5****: Comparison of AUC values for CL predicting sPTB < 37 weeks.*

| Reference | GA at mx | N | N sPTB | AUC |
|---|---|---|---|---|
| Current study | $18^{+0}$ - $22^{+0}$ | 1002 | 25 | 0.59 (0.45– 0.74) |
| Seravalli et al[39] | $24^{+0}$ - $27^{+6}$ | 1548 | 27 | 0.62 (0.50–0.74) |
| Kuusela et al[40] | $18^{+0}$ - $20^{+6}$ | 11072 | 585 | 0.60 (0.57–0.63) |
| Van der Ven et al[4] | $16^{+0}$ - $21^{+6}$ | 11943 | 464 | 0.61 (0.57-0.64) - NP<br>0.56 (0.52-0.60) - MP |
| Wikström et al[41] | $21^{+0}$ - $23^{+6}$ | 5582 | 110 – NP<br>59 - MP | 0.66 (0.60 – 0.71) – NP<br>0.53 (0.46 – 0.61) - MP |
| Gudicha et al[42] | $20^{+0}$ - $23^{+6}$ | 7336 | 1147 | 0.72 (0.69 – 0.75) |
| Thain et al[43] | $18^{+0}$ - $22^{+0}$ | 926 | 39 | 0.61 |
| Celik et al[44]<br>(this study excluded CL < 15 mm) | $20^{+0}$ - $24^{+6}$ | 58807 | 2216 | 0.617 |

GA: gestational age; sPTB: spontaneous preterm birth; AUC: area under the ROC curve; NP: nulliparous women; MP: multiparous women.

***Supplementary Table 6****: List of study sites, number of investigators at each site, number of women enrolled and number of women who completed the study, and country of the site.*

| Site No. | Site Name | # Investigators | # enrolled women | # completed women | Country |
|---|---|---|---|---|---|
| 1a | University Hospital Zurich USZ, Department of Obstetrics and Gynecology | 3 | 138 | 114 | CH |
| 1 | FMH Spezialarzt für Gynäkologie und Geburtshilfe - Satellite Site of USZ | 2 | 161 | 146 | CH |
| 1b | Frauengesundheitszentrum Zürich - Satellite Site of USZ | 1 | 11 | 9 | CH |
| 3 | Kantonsspital Aarau, Frauenklinik | 9 | 45 | 41 | CH |
| 4 | Kantonsspital Baden, Frauenklinik Geburtshilfe / Pränataldiagnositk | 2 | 81 | 76 | CH |
| 5 | Kantonsspital Frauenfeld, Frauenklinik | 2 | 183 | 175 | CH |
| 6 | Kantonsspital Luzern, Neue Frauenklinik | 4 | 90 | 81 | CH |
| 7 | Universitätsspital Basel, Klinik für Geburtshilfe und Schwangerenmedizin | 5 | 197 | 178 | CH |
| 8 | Frauenklinik, Stadtspital Triemli Zürich | 5 | 28 | 28 | CH |
| 9 | Kantonsspital St. Gallen, Frauenklinik | 3 | 46 | 39 | CH |
| 10 | Hôpitaux Universitaires Genève (HUG) | 5 | 50 | 45 | CH |
| 11 | Kantonsspital Münsterlingen | 2 | 25 | 23 | CH |
| 12 | Kantonsspital Winterthur | 1 | 9 | 8 | CH |
| 13 | Zentrum Zollikon | 1 | 2 | 2 | CH |
| 2 | University Hospital Leuven, Department of Obstetrics and Gynecology | 2 | 39 | 37 | BE |

### Supplementary Figures

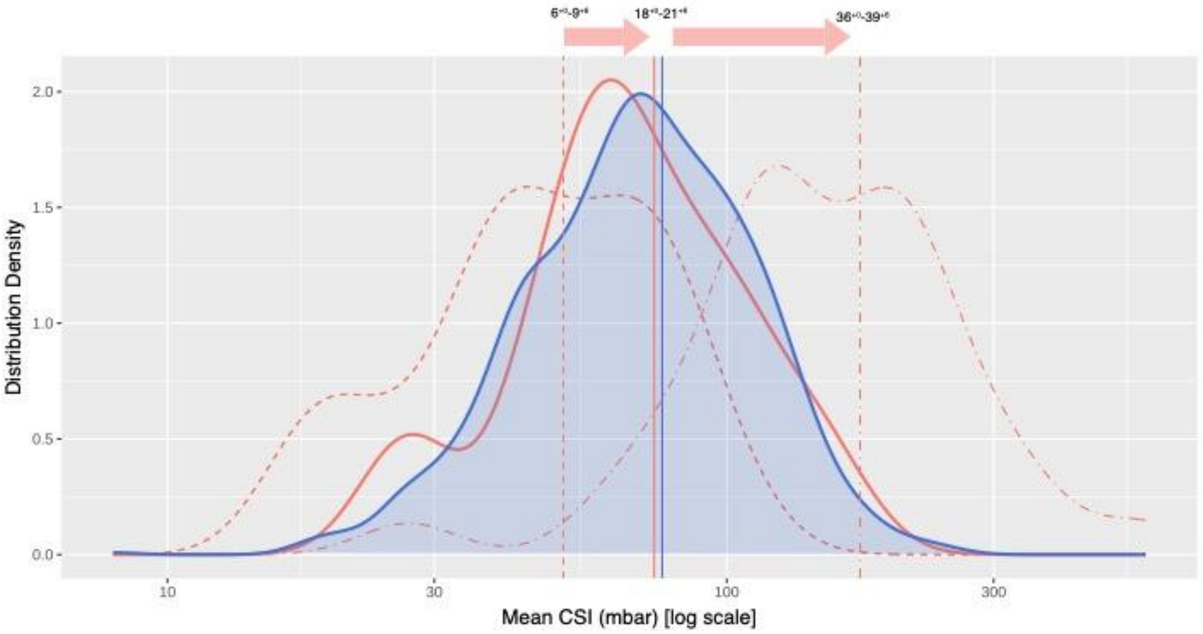


***Supplementary Figure 1:*** *Comparison of CSI log normal probability density distribution at $18^{+0}$ – $21^{+6}$ weeks for women with a singleton gestation who delivered at term, between the current study (blue, n = 950) and from Badir et al [1] (pink, n=42). Additional distributions are shown from the previous study early in the pregnancy at weeks $6^{+0}$ – $9^{+6}$ (dashed line) and later at weeks $36^{+0}$ – $39^{+6}$, (dashed-dotted line) illustrating the dynamic behavior of cervical stiffness throughout gestation. Vertical lines indicate the distribution medians.*


[1] Badir S, Mazza E, Zimmermann R, Bajka M. Cervical softening occurs early in pregnancy: characterization of cervical stiffness in 100 healthy women using the aspiration technique. Prenat Diagn. 2013;33(8):737-741. doi:10.1002/pd.4116

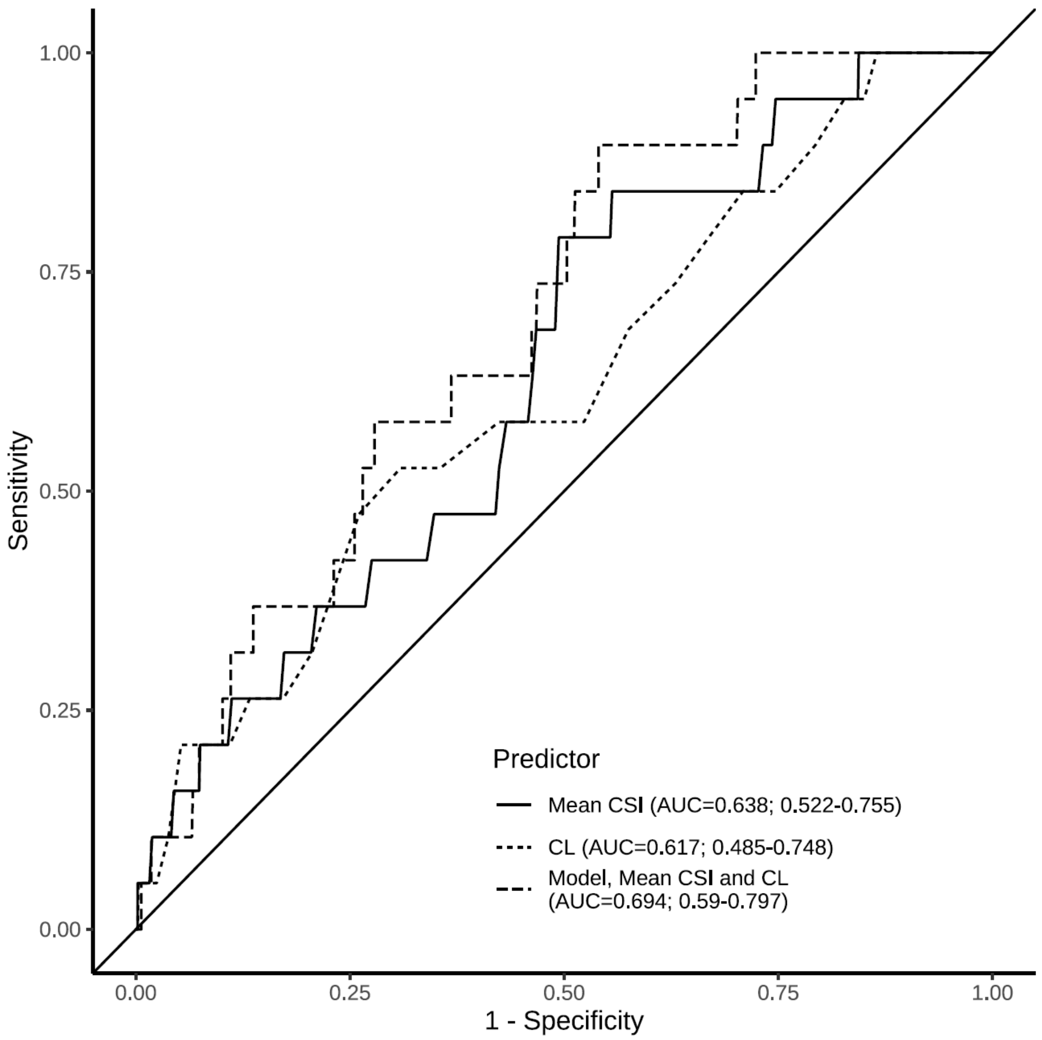


***Supplementary Figure 2:*** *ROC curves for spontaneous preterm birth prediction in women with a singleton gestation. Areas under the ROC curves (AUC) are: mean CSI 0.638 (95% CI, 0.522 – 0.755), CL 0.617 (95% CI, 0.485 – 0.748), model including CSI and CL 0.694 (95% CI 0.590 – 0.797).*
